# The Role of Children in SARS-CoV-2 Variant of Concerns Transmission within Households: A Meta-analysis

**DOI:** 10.1101/2022.07.21.22277914

**Authors:** Yanshan Zhu, Yao Xia, Janessa Pickering, Asha C. Bowen, Kirsty R. Short

## Abstract

**Background:** Meta-analysis and single site studies have established that children are less infectious within a household when positive for ancestral SARS-CoV-2. In addition, children appear less susceptible to infection when exposed to ancestral SARS-CoV-2 within a household. The emergence of SARS-CoV-2 variants of concern (VOC) has been associated with an increased number of pediatric infections worldwide. However, the role of children in the household transmission of VOCs, relative to the ancestral virus, remains unclear.

**Methods:** Here, we perform a meta-analysis of the role of children in the household transmission of both ancestral SARS-CoV-2 and SARS-CoV-2 VOCs. Results: Unlike the ancestral virus, children infected with VOCs spread SARS-CoV-2 to an equivalent number of household contacts as infected adults. Similarly, unlike the ancestral virus, children within a household were equally as likely as adults to acquire SARS-CoV-2 from an infected family member. Interestingly, this same observation was noted when unvaccinated children exposed to VOCs were compared to unvaccinated adults exposed to VOCs.

**Conclusions:** Together, these data suggest that the emergence of VOCs were associated with a fundamental shift in the epidemiology of SARS-CoV-2. This is unlikely to solely be the result of age-dependent differences in vaccination during the VOCs period and instead may reflect virus evolution over the course of the pandemic.

**summary:** The role of children in the household transmission of VOCs, relative to the ancestral virus, remains unclear. Using a meta-analysis we show that the emergence of VOCs were associated with a fundamental shift in the epidemiology of SARS-CoV-2

## Introduction

In the first six months of the Severe acute respiratory syndrome coronavirus 2 (SARS□CoV□2) pandemic there were numerous household transmission studies suggesting that compared to adults, children were less susceptible to SARS-CoV-2 infection and less likely to transmit the virus [1]. These findings were echoed in studies outside of households where the infection rate of SARS-CoV-2 amongst children less than 10 years old was significantly lower than that of adults [2]. However, since August 2020, the continuous emergence of new variants of SARS-CoV-2 has raised questions about a fundamental shift in the epidemiology of SARS-CoV-2[3].

Globally, there has been at least three peaks corresponding to the prevalence of variants of concern (VOCs) Alpha (or Beta/Gamma), Delta and Omicron [3, 4]. During these waves there has been speculation that children have become more susceptible to SARS-CoV-2 infection and more infectious once they contract the virus. For example, during the Delta wave in Singapore children (aged 0-11 years) were significantly more likely to transmit and acquire SARS-CoV-2 in a household compared to young adults (18-29 years) [5]. Similarly, during the Omicron wave in the USA the secondary attack rates (SAR) were consistently high across household contact and index age groups, including those aged 0-4 years old [6].

Despite these data from single site studies, meta-analysis on the role of children in the spread of VOCs (relative to the ancestral virus) are generally lacking, with studies often focused on the ancestral virus [7] or not differentiating between data collected during the pre- and post-VOCs period [8]. Where pre-VOCs and post-VOCs based studies have been differentiated, data suggests an increased role for children in the household transmission of VOCs [7]. However, such analysis remains confounded by the fact that globally, adults have been prioritized for vaccination [9]. Vaccination campaigns for COVID-19 were largely rolled out from December 2020 onwards but with a primary focus on vaccinating individuals over the age of 18 years. Vaccination of children 5-11 years of age was not approved by the U.S. Food and Drug Administration (FDA) until October 2021[10]. Pediatric vaccination rates remain consistently lower than those of adults [11], with FDA approval for vaccination in children 6 months to under five years of age only approved in June 2022. Vaccination significantly reduces the household transmission of SARS-CoV-2 [12]. As a result, it is difficult to ascertain if any observed epidemiological changes in the demographics of viral transmission over time are the result of a fundamental change in the virus over time or if a potential increase in pediatric infections and transmission is simply indicative of the lower vaccination rate amongst children.

To assess what effect the SARS-CoV-2 variants of concern (VOCs) have on children in terms of infectiousness and susceptibility to SARS-CoV-2 infection within a household, here we perform a meta-analysis comparing pediatric SARS-CoV-2 transmission during the pre and post VOCs period.

## Methods

### Case definitions

We adapted the WHO household transmission investigation protocol for coronavirus disease 2019 (COVID-19) [13]. A household transmission was defined as a group of people (2 or more) living in the same residence in whom two or more positive SARS-CoV-2 cases subsequently occurred during the follow-up visits within 28 days of the index. An index case was defined as the first case of laboratory-confirmed COVID-19 in the same household. A secondary case was defined as a known household contact of the confirmed infected index who has been tested positive for SARS-CoV-2 during the follow up period. A household contact was defined as a person who has cohabited with the index case in a defined period of time. In this context, the secondary attack rate is a measure of the frequency of secondary infections of COVID-19 among household contacts in a defined period of time, as determined by a positive COVID-19 result. Adults were defined as individuals ≥18 years old whilst children were defined as individuals below 18 years old.

### Classification of SARS-CoV-2 Variants of Concern by study period

Studies were classified as pertaining to the ancestral virus or a VOC based on available genotype data and/or the timing of the study period. Specifically, studies where SARS-CoV-2 genotype was not documented, and the index case identification period was prior to January 1, 2021, were defined as the period of ancestral virus predominance. Studies where data was collected between January 1, 2021 and June 30, 2022 were categorised as VOCs period.

### Vaccination status

In investigating the effect of vaccination on transmission, only studies reporting vaccination status of household contacts were included. Vaccinated individuals were defined as those who had received at least one dose of a SARS-CoV-2 vaccine.

### Search strategy and eligibility criteria

Literature searching was performed in accordance with Preferred Reporting Items for Systematic Reviews and Meta-Analyses (PRISMA) statement[14]. Our original systematic review screened literatures through August 24, 2020[1], therefore in this study, publications available between August 25, 2020, and June 30, 2022, were accessed from PubMed, Covid MEDLINE, Embase, and Web of Science, using the search term: (“COVID-19” OR “SARS-CoV-2” OR “variant”) AND (“household transmission” OR “family cluster” OR “household contact”) OR (“transmissibility” OR “attack rate”) OR (“vaccination” OR “attack rate”) with no language or location restrictions. Given the role of preprints in timely dissemination of research findings during the COVID-19 pandemic, we also conducted searches of the medRxiv and bioRxiv servers using the search term (“COVID-19” OR “SARS-CoV-2”) AND (“household transmission” OR “secondary attack rate”) for the posted articles. Investigator YZ developed the initial search strategy, which was then validated by YX and JP. Additional relevant articles from the reference sections were also reviewed. Studies that were duplicate publications, modeling studies, case reports and/or reviews were excluded (Fig. 1).

**Fig. 1.**
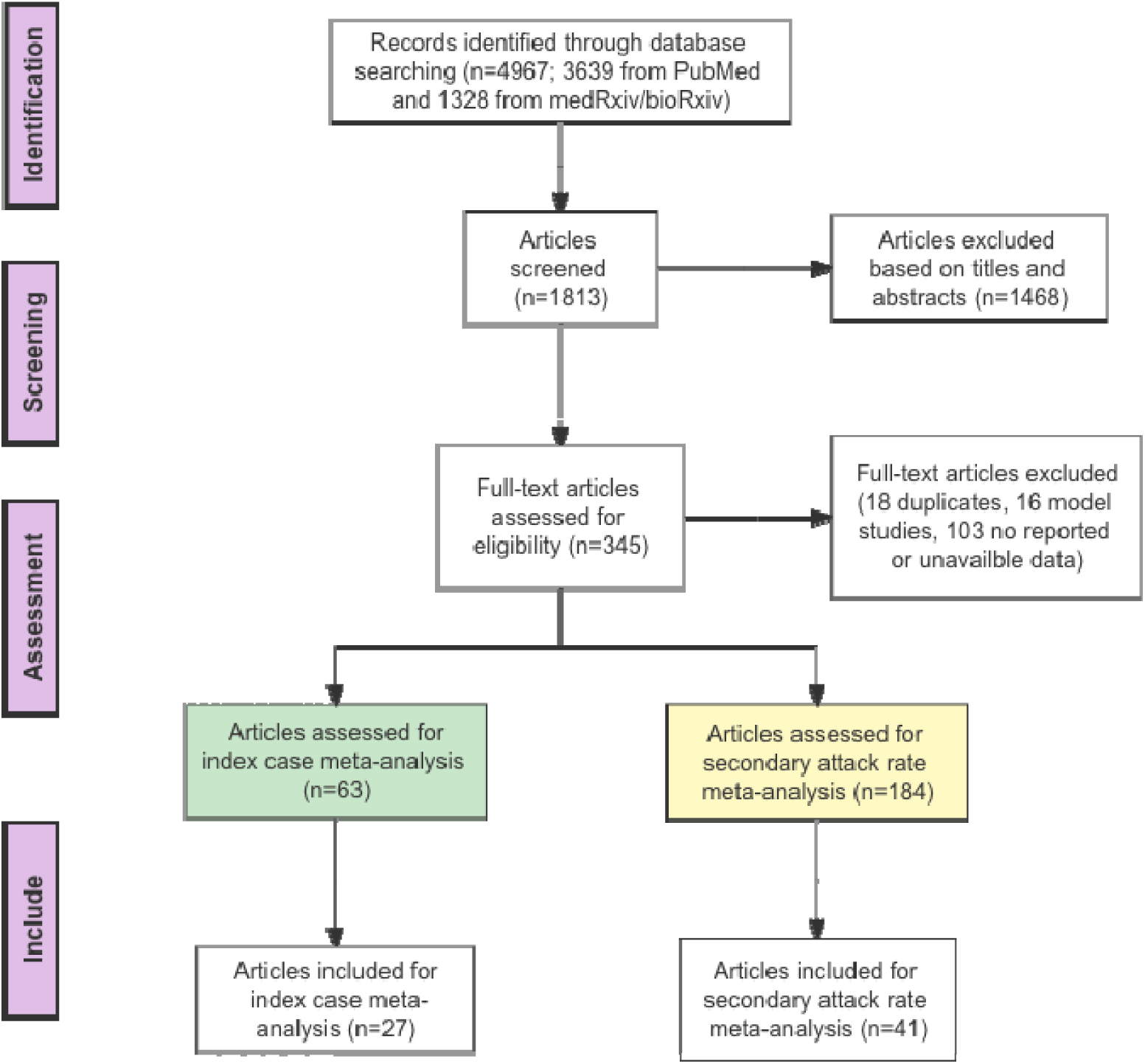
Preferred Reporting Items for Systematic Reviews and Meta-analysis (PRISMA) flow diagram.

### Statistical analysis

We assessed the infectiousness and susceptibility of children and adults to SARS CoV-2 infection during different time periods by pooling all data deemed eligible by the above selection criteria. Susceptibility to SARS CoV-2 VOCs infection was estimated by pooling the secondary attack rates (SARs) for household contacts. We estimated the Relative Risk (RR) for SARS CoV-2 household secondary infection stratified by the age of index cases, the age of household contacts, and the vaccination status of household contacts for each study. We then used Generalized Linear Mixed Models[15] to estimate pooled RRs along with corresponding 95% confidence intervals. As only observational studies were used, we used a random effects model, equalizing the weight of the studies to the pooled estimate. Where relevant, we stratified the analysis by pre-specified characteristics including the characteristics of index cases and contacts. Heterogeneity between studies was evaluated using the I^2^ statistic test. A threshold of I^2^ > 50% was used as indicating statistically significant heterogeneity. All summary analyses and meta-analysis were performed using R studio software (version 3.6.1).

## Results

A total of 4967 studies (3936 publications and 1328 preprint articles) were initially identified from the literature search. Among these studies 1468 were excluded based on title and abstract. After further evaluation, 345 studies were eligible for full text review and 48 studies met the inclusion criteria, of which 27 contained eligible data included for the index case meta-analysis and 41 new studies combining with 11 studies from our previous review [1] for secondary attack rate meta-analysis (Table S1). Specially, twenty studies involving 12 countries (USA, South Africa, Netherlands, UK, Norway, Israel, Germany, Denmark, Singapore, Japan, India and Thailand) assessed the secondary attack rate during the period of VOCs dominance. Twenty-seven studies from 16 countries (Brazil, China, India, Greece, Japan, USA, UK, Spain, Bosnia and Herzegovina, Malaysia, Netherlands, Israel, Australia, Norway, Singapore and Canada) were subjected to analysis for secondary attack rate in the period defined as pre-VOCs (Table S1).

### Children were less infectious within households during the period that the ancestral virus was dominant

Fourteen studies were identified that defined the age of the index case and the SAR in the household during the time period in which the ancestral virus was dominant (until 1 January 2021). Another 14 studies were identified that defined the age of the index case and the SAR in the household during the time period in which VOCs were dominant. An increasing trend of estimated SAR overtime is shown in Fig. 2. During the time period when the ancestral virus was dominant (before 1^st^ January 2021), a pediatric index case was associated with a significantly lower secondary attack rate compared to an adult index case (RR= 0.61; 95% CI, 0.47-0.80, Fig. S1). In contrast, there was no significant difference with SAR (RR = 0.98; 95% CI: 0.85-1.13) between a pediatric index case and an adult index case during the VOCs period (Fig. S1). The role of younger children (<12 years old) in transmitting a VOC within the household was then examined by 8 observational studies with no significant heterogeneity (I^2^= 19%, P =0.29), which involve pediatric index cases of different ages. The secondary attack rates caused by a younger pediatric index post-VOCs were higher than rates attributable to an older child index case, in which we found an estimated 46% significant increase in SAR among household contacts (Fig. S2). Together, these data suggest that children, especially younger children, were more infectious in the family during the period of VOCs dominance.

**Fig. 2.**
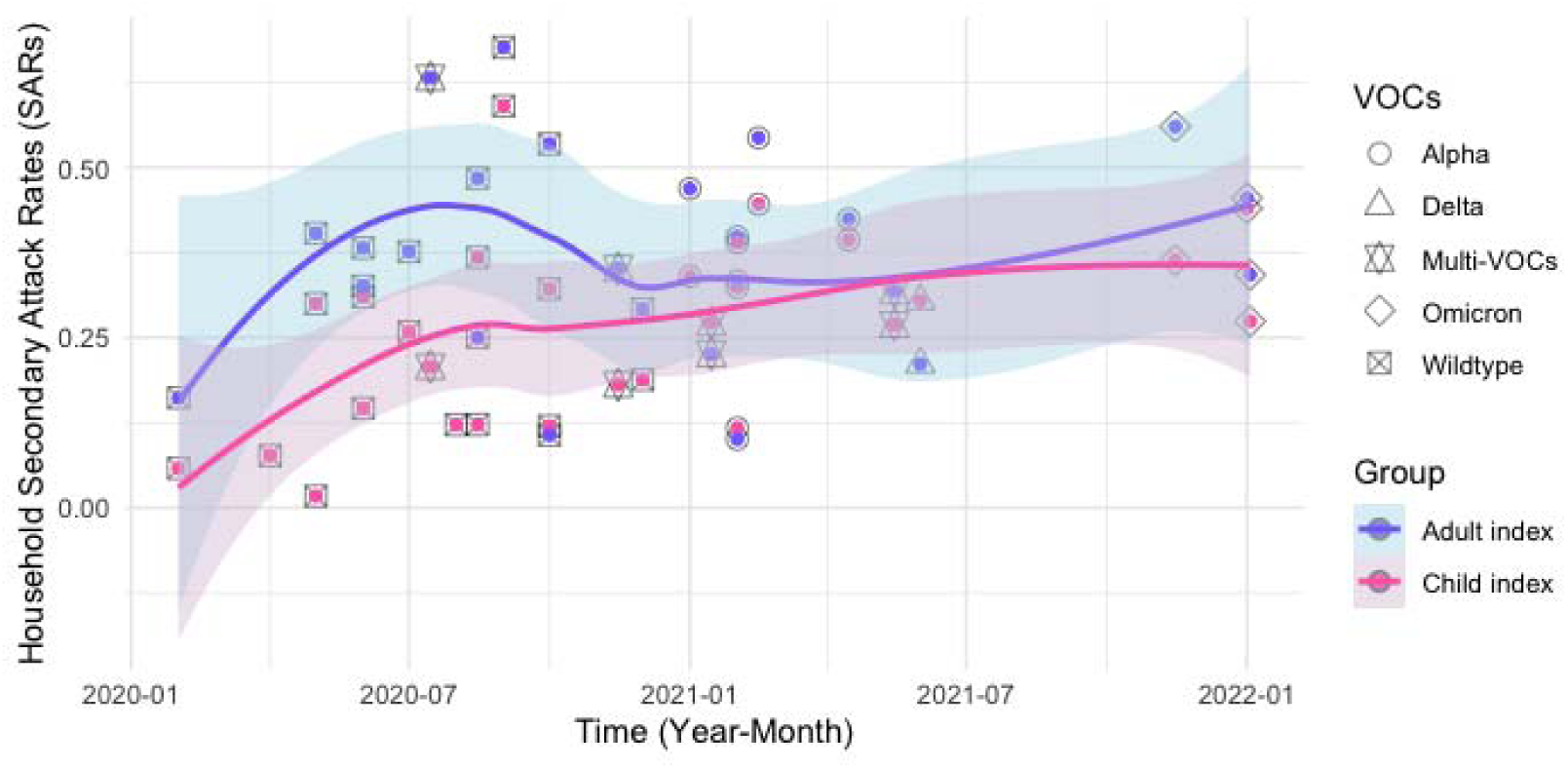
Time pattern for the secondary attack rate from a child index and an infected adult index in household SARS-CoV-2 transmission and smoothed curve of SARs by month during the study period. Each time point is based on the midpoint of study period in each study. Multi-VOCs points represent studies with 2 or more predominant variants.

### Children have a higher secondary attack rate in household transmission of SARS-CoV-2 during the period of VOCs dominance

To determine the susceptibility of children to household SARS-CoV-2 infections, the secondary attack rate in the household contacts was assessed in 27 pre-VOC studies and 20 VOC studies. The increasing trends of secondary attack rate among children and adults contacts in household SARS-CoV-2 transmission was associated with the growing dominance of SARS-CoV-2 VOCs since 2021 (Fig. 3). The random effects model results suggest that children were statistically less likely to acquire ancestral SARS-CoV-2 (SAR=18%, 95% CI: 0.12-0.26) than VOCs (SAR=31%, 95% CI: 0.24-0.38, Fig. S3), and subgroup difference test showed *P*<0.01. In contrast, before VOCs were dominant, the average pooled SAR of adult (SAR=29%, 95% CI: 0.23-0.39) was similar with those during the VOC period (SAR=31%, 95% CI: 0.26-0.37) (*P*=0.64, Fig. S4). As shown in Fig. 3B, household SARs were higher for VOCs among children contacts compared to the ancestral virus and equivalent to those among their adult family members.

**Fig. 3.**
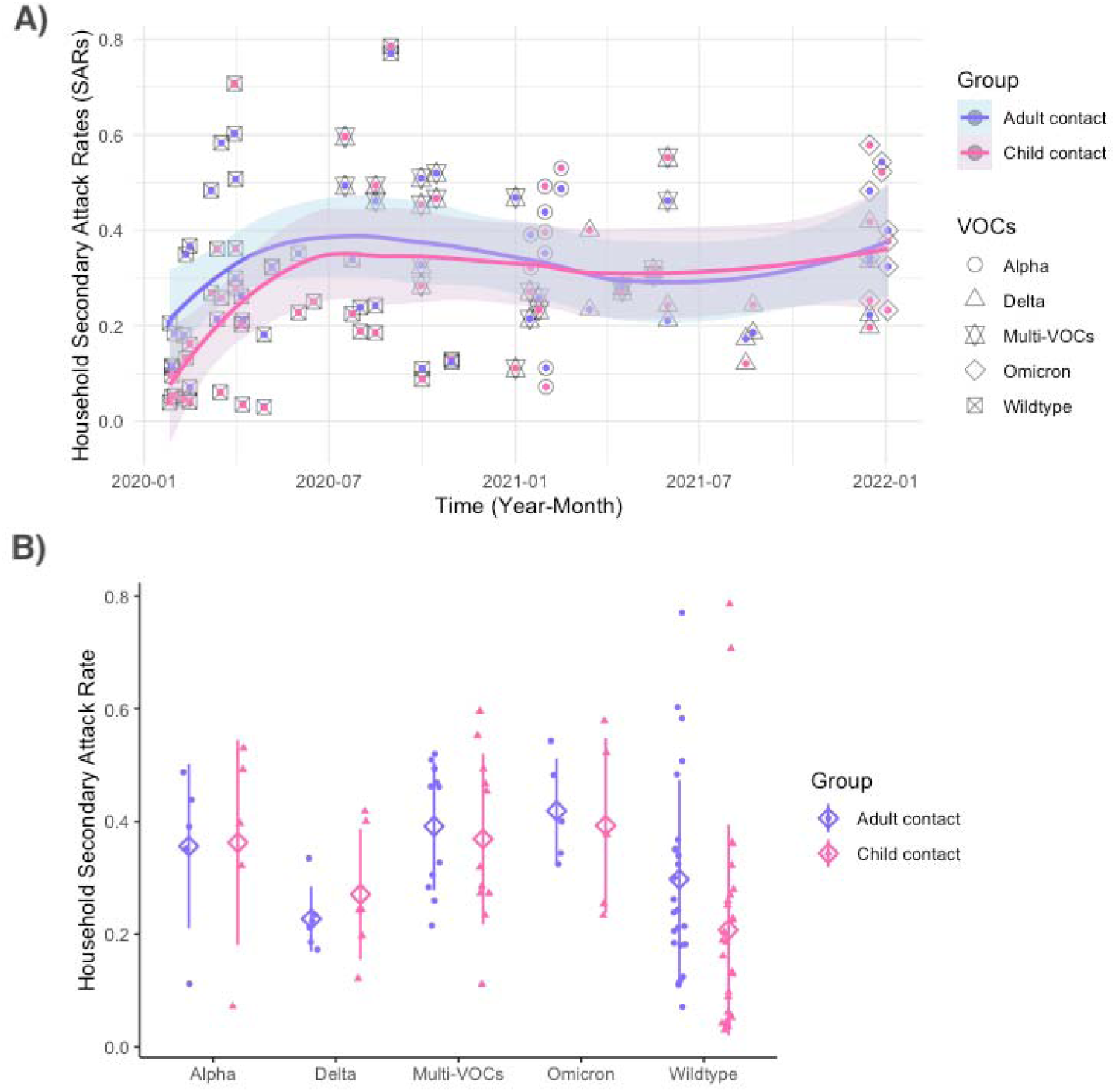
A) Time pattern for the secondary attack rate among children and adults contacts in household SARS-CoV-2 transmission and smoothed curve of secondary attack rates by month during the study period. Each time point is based on the midpoint of study period in each study. Multi-VOCs points represent studies with 2 or more predominant variants. B) The secondary attack rate in household SARS-CoV-2 transmission among children and adults contact stratified by different VOCs.

Although we observed significant heterogeneity between the included studies, in a subset analysis where additional information was provided on the age of the pediatric contact, younger children (<12 years) were no more or less susceptible to infection compared with older children (≥12 years) during pre-VOCs period; RR = 0.77 (95% CI, 0.59-1.02, Fig. 4). This is consistent with our prior analysis of the SAR in children and adults during the first year of COVID-19 pandemic [1]. However, the period of VOCs shows a different scenario, in which there was an estimated 46% statistically significant increase in SAR among younger children household contacts. RR = 1.46 (95% CI, 1.10-1.94, Fig. 4). In addition, our findings show that compared to older children, estimated risks of younger children acquiring SARS-CoV-2 was significantly different in the two periods (*P*<0.01), although the heterogeneity among the observational studies during the post VOCs was high (I^2^ = 97%, P < 0.01).

**Fig. 4.**
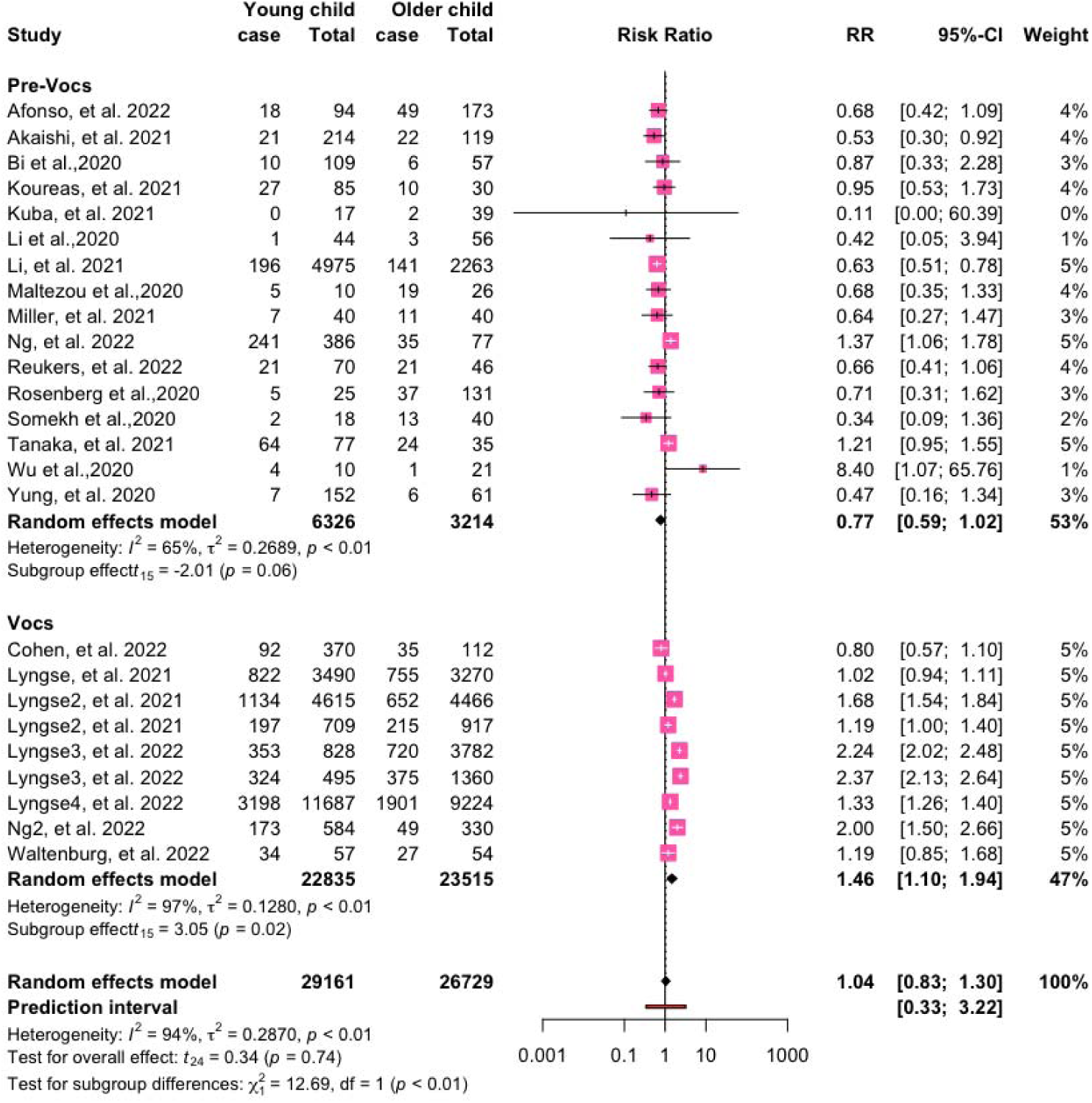
Relative risk (RR) for the secondary attack rate of younger and older children contacts in household severe acute respiratory syndrome coronavirus 2 (SARS-CoV-2) transmission stratified by the pre and post VOCs period. Cases describe the number of SARS-CoV-2 positive individuals identified in the study. Abbreviation: CI, confidence interval.

The above studies were classified as the pre- or post-VOCs period based on the time at which the data was collected. To confirm the same results were obtained when specific virus genotyping was performed, the same analysis was performed with a subset of studies where the specific VOC was determined. Consistent with our prior results[1], children were significantly less likely to acquire the ancestral virus in the household compared with adults (Fig. S5). In contrast, the risk of children being infected with Alpha, Delta or Omicron was not significantly different from exposed adult household contacts (Fig. S5).

The above data suggest that children were more infectious (Fig. 2, Fig. S1) and more susceptible to infection (Fig. 3, Fig. S3) during the period at which VOCs were dominant. However, this was also the period at which vaccination amongst adults became widespread. To determine if age-dependent differences in vaccination affected these data we examined SARs by vaccination status (during the VOC period only) of household contacts regardless of vaccination status or age of the index cases. Only nine studies from Netherlands, USA, Norway, Israel, Denmark, Singapore, Japan, and UK, reported the effectiveness of vaccination against secondary transmission of SARS-CoV-2 within the household. Consistent with previous studies [16], the estimated SAR was higher for unvaccinated adult contacts than fully or partially vaccinated adults, RR = 1.78 (95% CI, 1.49-2.13) with heterogeneity (I^2^ = 78%, P < 0.01, Fig. 5). These data demonstrate that vaccination can affect SARs within the household. To address this issue in the context of age-dependent differences in vaccination and transmission, we analysed a subset of studies that investigated the SAR in unvaccinated children and unvaccinated adults collected during the period of VOC dominance. In the absence of vaccination there was no difference in the SAR between adults and children (RR = 0.91 (95% CI, 0.78-1.07; Fig. 5). This is consistent with our prior analysis of the SAR in children and adults (Fig. S5).

**Fig. 5.**
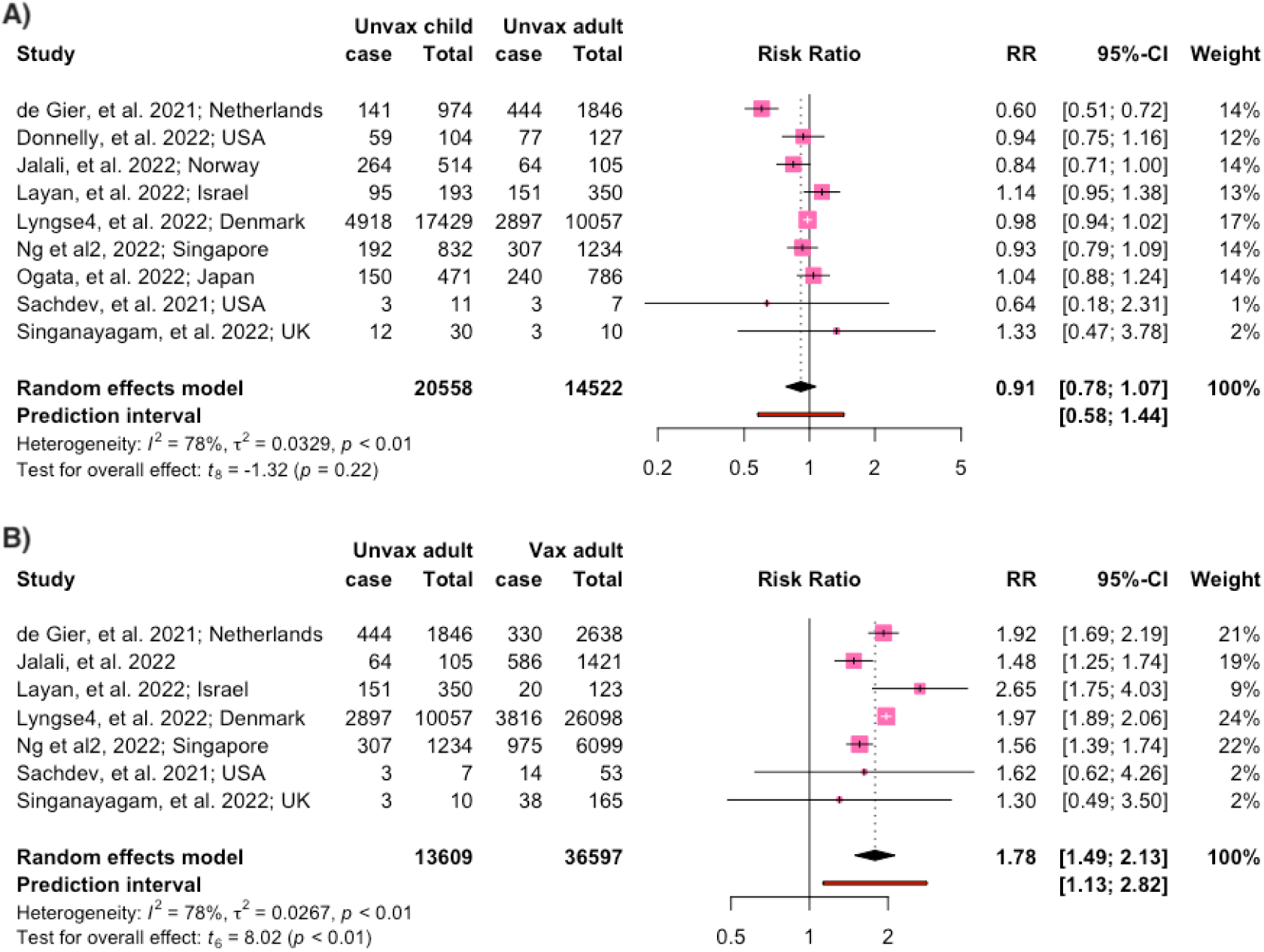
Relative risk (RR) for the SARs of child contact and adult contact in household SARS-CoV-2 VOCs transmission Stratified by vaccination status. Cases describe the number of SARS-CoV-2 positive individuals identified in the study. Abbreviation: CI, confidence interval. Vaccinated adults are defined as having received at least one dose of vaccine

## Discussion

In the early stages of the COVID-19 pandemic children did not appear to play a significant role in the household transmission of SARS-CoV-2 [1]. Data presented here suggest that this has shifted throughout the course of the pandemic, and in particular since the emergence of VOCs in the community.

The increased role for children in the household transmission of SARS-Cov-2 during the post-VOC period adds weight to the importance of COVID-19 vaccination programs in children, including those now beginning in <5 years. This remains a contested issue with many parents finding, in light of the typically mild disease children experience when infected with SARS-CoV-2, it difficult to make an informed risk-benefit assessment regarding pediatric vaccination [17]. Whilst the slower rollout of pediatric COVID-19 vaccination has precluded detailed assessments of the precise effect of pediatric vaccination on household SARS-CoV-2 transmission, it is promising that vaccination reduced the risk of infection among cohabitating adults/teenagers [18] and that the person-to-person probability of SARS-CoV-2 transmission between two vaccinated adults/teenagers was 4% (compared to the 61% observed between unvaccinated household members) [18]. These data may suggest that pediatric COVID-19 vaccination during the VOC period will not only reduce the risk of severe disease in the child but may also play an important role in reducing household transmission of the virus.

It remains to be determined if the data shown herein can be translated to scenarios outside of the home (e.g., SARS-CoV-2 transmission in the school settings). However, even if this should be the case, it is important not to interpret these data as a rationale for re-introducing or continuing school closures. In contrast to the early stages of the COVID-19 pandemic, high adult vaccination rates and the increased availability of pediatric vaccination, combined with a global decline in the severity of SARS-CoV-2 cases and improved disease prevention measures (e.g., ventilation, mask use) represents an opportunity for continued face-to-face schooling. However, it is clear from these data that public health decisions such as these need to be derived from data with the current circulating SARS-CoV-2 variants, and not the ancestral virus, in order to most accurately represent the present situation. Prioritising business as usual for all domains of society and layering public health measures on top of these has become operational in most countries learning to live with COVID-19, and this household transmission data adds weight to the importance of this. Whilst children are long thought to be vectors of high viral transmission e.g., influenza, RSV, SARS-CoV-2 still does not follow this trend. Children are equally as likely as adults in the post-VOC era to transmit SARS-CoV-2, but no more likely than adults, unlike in the seasonal influenza or RSV patterns [19, 20].

This study has also provided a new insight into the possible causes of increased VOC transmission amongst children relative to the ancestral virus. It is possible that these data represent differential COVID-19 vaccination rates between children and adults, given the role that vaccination can play in preventing the household transmission of SARS-CoV-2[12]. However, such a hypothesis would suggest that a comparison of household transmission amongst unvaccinated adults and children during the VOC period would show a pattern akin to that of the ancestral virus (i.e., an age-dependence difference in susceptibility to infection and infectiousness within a household). Instead, amongst unvaccinated individuals during the VOC period there was no age-dependent differences in household SARS-CoV-2 transmission. These data are consistent with different adult and pediatric vaccination playing a minimal role in the changing epidemiology of SARS-CoV-2 over the course of the pandemic. Instead, these data may suggest that the evolution of virus over time has resulted in an increased role for children in viral transmission. Indeed, we have recently shown that the ancestral SARS-CoV-2, but not Omicron, is less efficient at replicating in the primary nasal epithelial cells of children [21] – which may have implications for how much virus a child versus an adult shed within the household. However, it does remain possible that the observed shifts in the epidemiology of SARS-CoV-2 over time represent changes in the virus in addition to age-dependent differences in both vaccination and infection. This represents an important area of ongoing research.

This study was subject to several limitations. Firstly, only a limited number of studies were available in the post-VOCs period that documented household SARS-CoV-2 transmission amongst unvaccinated adults and children. Moreover, the often-mild nature of Omicron infection may have meant that the SAR in Omicron studies were underestimated. Nevertheless, these data provide the first comprehensive insight into the shifting role of children in virus transmission over the course of the SARS-CoV-2 pandemic.

## Data Availability

All data produced in the present work are contained in the manuscript

## Author contributions

YZ: conceptualization, methodology, data analysis, writing—original draft preparation; YX: methodology, data analysis, writing—reviewing and editing; JP: data analysis, writing—original draft preparation; AB: conceptualization. Writing—reviewing and editing; KS: conceptualization, methodology, writing—original draft preparation. All authors read and approved the final manuscript.

## Funding

This work is supported by the National Health and Medical Research Council investigator grant 2007919 to K.R.S..

## Declarations

### Competing interests

KRS is a consultant for Sanofi, Roche and NovoNordisk. The opinions and data presented in this manuscript are of the authors and are independent of these relationships.

### Ethical approval

This is a meta-analysis study. The University of Queensland Research Ethics Committee has confirmed that no ethical approval is required.

### Consent to participate

Not applicable.

## Notes

### Competing Interest Statement

The authors have declared no competing interest.

