## Supplemental figures for "The Role of Children in SARS-CoV-2 Variant of Concerns Transmission within Households: A Meta-analysis"

Yanshan Zhu^1#^, Yao Xia^2,3#^, Janessa Pickering^4^, Asha C. Bowen^4,5^ & [Kirsty R. Short](about:blank" \l "xce9e4f69)^1,6*^

**Supplementary Figures**

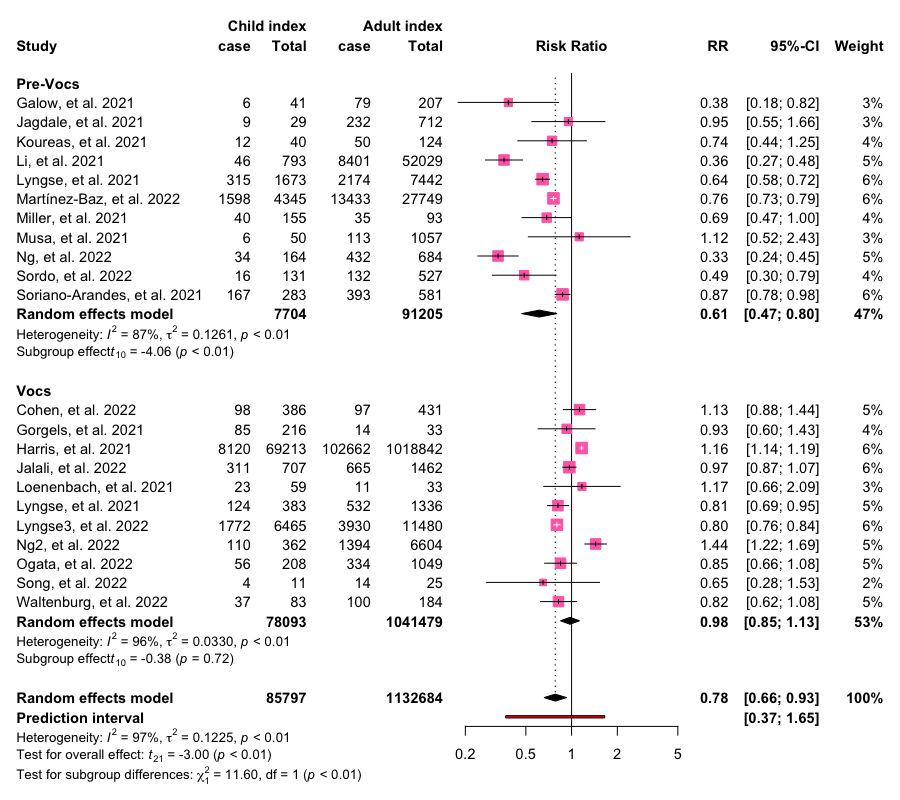

**Fig. S1** Relative risk (RR) for the secondary attack rate of household members when an adult or a child was identified as the index case. Cases describe the number of SARS-CoV-2 secondary infection identified in the study. Abbreviation: CI, confidence interval.

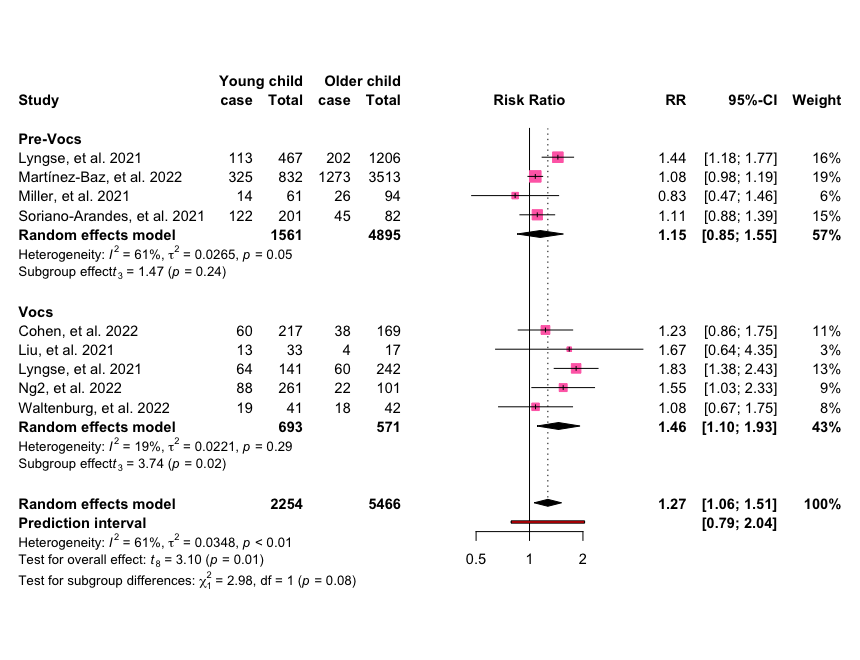

**Fig. S2** Relative risk (RR) of the secondary attack rate of household members when a younger or older child was identified as the index case. Cases describe the number of SARS-CoV-2 secondary infection identified in the study. Abbreviation: CI, confidence interval.

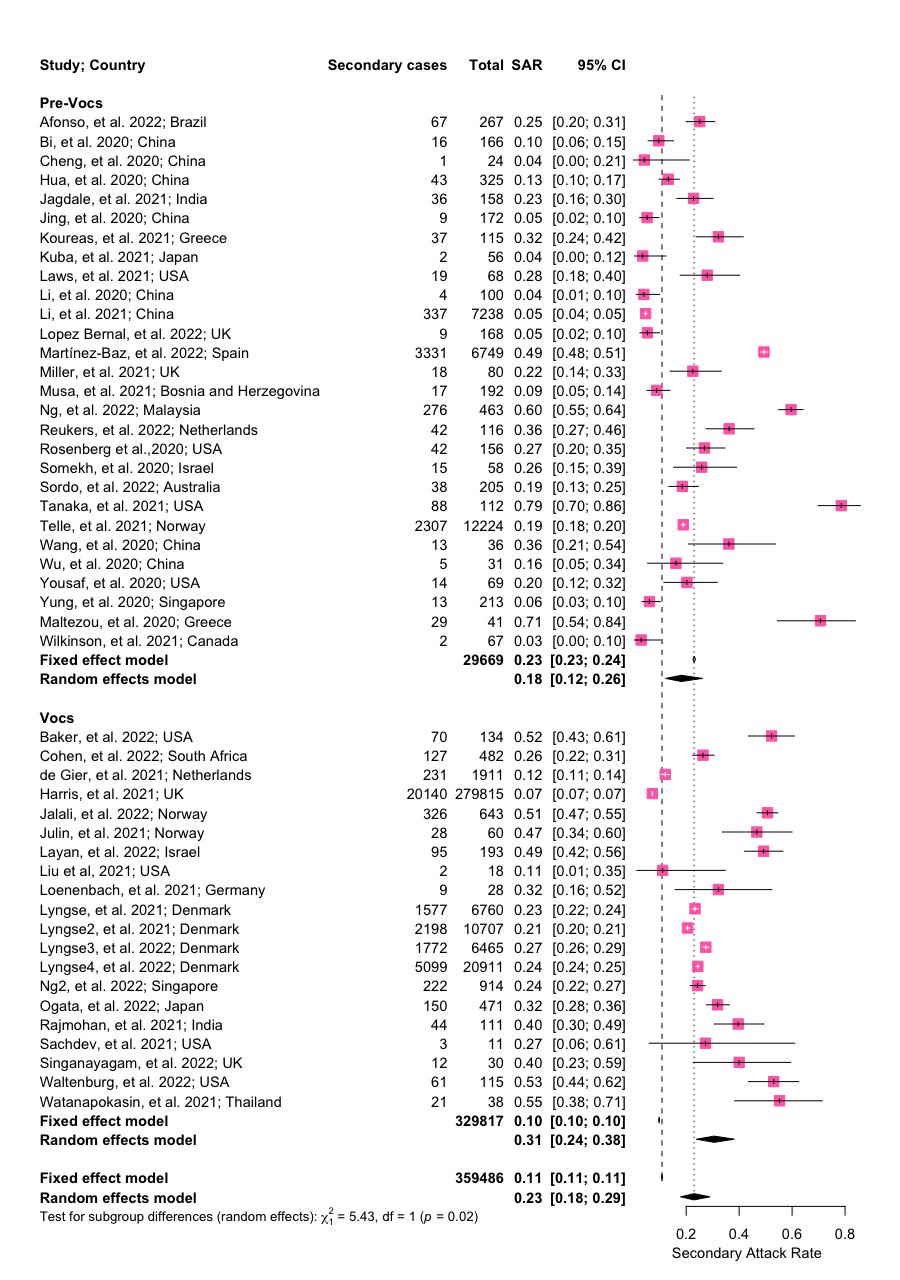

**Fig. S3** The secondary attack rate of children contact in household severe acute respiratory syndrome coronavirus 2 (SARS-CoV-2) transmission stratified by the pre and post VOCs period.

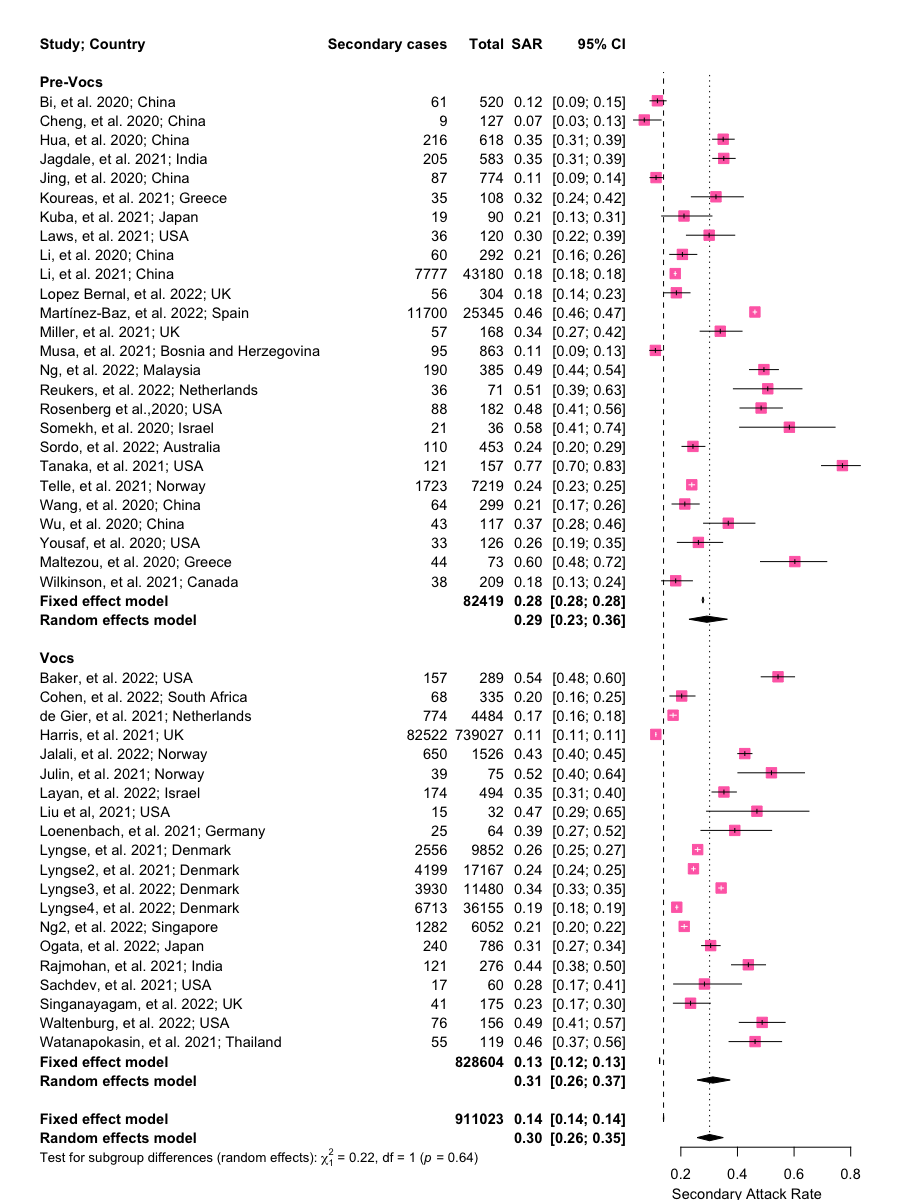

**Fig. S4** The secondary attack rate of adults contact in household severe acute respiratory syndrome coronavirus 2 (SARS-CoV-2) transmission stratified by the pre and post VOCs period.

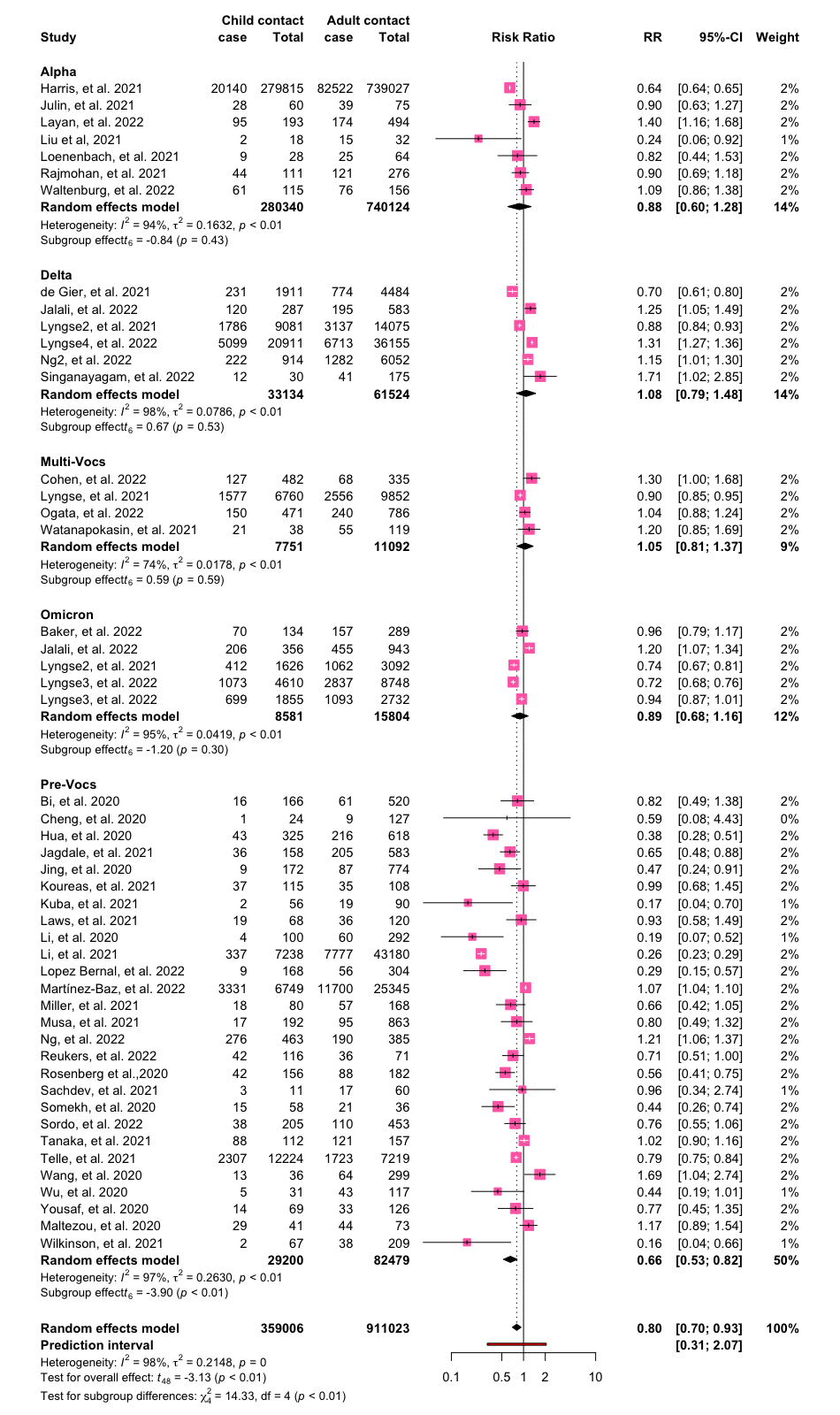

**Fig. S5** Relative risk (RR) of child contact and adult contact in household transmission of Alpha, Delta, Multi-VOCs or Omicron variants compare with the Pre-VOCs period. Cases describe the number of SARS-CoV-2 secondary infection identified in the study. Abbreviation: CI, confidence interval.
